# Scaling COVID-19 against inequalities: should the policy response consistently match the mortality challenge?

**DOI:** 10.1101/2020.05.04.20090761

**Authors:** Gerry McCartney, Alastair H. Leyland, David Walsh, Ruth Dundas

## Abstract

**Background:** The mortality impact of COVID-19 has thus far been described in terms of crude death counts. We aimed to calibrate the scale of the modelled mortality impact of COVID-19 using age-standardised mortality rates and life expectancy contribution against other, socially-determined, causes of death in order to inform governments and the public.

**Methods:** We compared mortality attributable to suicide, drug poisoning and socioeconomic inequality with estimates of mortality from an infectious disease model of COVID-19. We calculated age-standardised mortality rates and life expectancy contributions for the UK and its constituent nations.

**Results:** Mortality from a fully unmitigated COVID-19 pandemic is estimated to be responsible for a negative life expectancy contribution of −5.96 years for the UK. This is reduced to −0.33 years in the fully mitigated scenario. The equivalent annual life expectancy contributions of suicide, drug poisoning and socioeconomic inequality-related deaths are −0.25, −0.20 and −3.51 years respectively. The negative impact of fully unmitigated COVID-19 on life expectancy is therefore equivalent to 24 years of suicide deaths, 30 years of drug poisoning deaths, and 1.7 years of inequality-related deaths for the UK.

**Conclusion:** Fully mitigating COVID-19 is estimated to prevent a loss of 5.63 years of life expectancy for the UK. Over 10 years there is a greater negative life expectancy contribution from inequality than around six unmitigated COVID-19 pandemics. To achieve long-term population health improvements it is therefore important to take this opportunity to introduce post-pandemic economic policies to ‘build back better’.

**Summary box:** *What is already known on this subject?:* COVID-19 has been modelled to create a substantial excess mortality in the UK, depending on the degree to which this is mitigated by social distancing measures. Best estimates of 510,000 and 20,000 crude deaths are predicted in unmitigated and fully mitigated scenarios respectively.

*What does this study add?:* We scale the mortality impact of the modelled COVID-19 on age-standardised mortality and life expectancy against suicide, drug poisonings and inequalities. The impact of COVID-19 on life expectancy is substantial (−5.96 years) if unmitigated, but over a decade the life expectancy impact of inequalities is around six times greater than even an unmitigated pandemic.

## Background

The COVID-19 pandemic has been tracked by daily counting of cumulative numbers of confirmed cases and deaths.^1^ The exponential growth in these numbers across countries has understandably created anxiety and action from public health agencies and governments internationally. Despite initial surveillance and reporting of COVID-19 following standard infectious disease epidemiologic methods, the subsequent reporting of COVID-19 mortality has largely focused on crude death counts, arguably not meeting the “rigorous standardisation and quality control of investigative methods [that] are essential in epidemiology”.^2^ A number of particular limitations in the data have prevented a sufficient understanding the true impact of the pandemic on mortality.

First, the counting of cases of COVID-19 within and between countries has been dependent on the case definition and the changes in that over time. At the beginning of the outbreak in Wuhan, China, cases were defined clinically before virological testing was available. Then, as testing became partially available, cases were defined as people with a travel history from China, or contact with a known case, and a positive virology test. Then cases were defined as people with a positive virology test irrespective of symptoms or history, although the availability of tests remained restricted.^3^ This is problematic for epidemiological surveillance, because limited availability of testing, and the self-limiting nature of the infection for many, meant that the case count underestimated the true incidence within the population. For COVID-19 deaths, the count was initially based on people who died within hospital who had a positive virological test. This is subsequently being extended in most countries to include coding of deaths based on clinical opinion in all settings. This raises a further issue because many deaths will occur in people who die with, rather than die of, COVID-19.

Deaths occur every day, and simply reporting the *cumulative* number of deaths for any particular cause, be that COVID-19 or anything else, will always reveal a rising trend. Other causes of death are not reported in this way. It is therefore difficult for the public and policymakers to understand how to interpret and compare these to other causes of death.

The COVID-19 deaths reported are crude deaths counts. They therefore do not take into account the size of the population at risk (as a crude rate does), nor the age and sex structure of the population, in particular how old the population is (as an age-sex-standardised rate does). In contrast, other causes of death such as cancers and heart disease, and deaths attributable to deprivation, poverty and other political and socioeconomic causes such as austerity, are usually measured as differences in such standardised rates, in years of Life Lost (YLL), or life expectancy contributions.^4^ Finally, the reported crude death counts also do not account for competing causes and how likely people dying from COVID-19 were to have died relatively soon from other causes.^5^ It is therefore difficult to assess the scale of the mortality risk of COVID-19 relative to the background mortality risk in the population.

These problems of interpretation are very important. If COVID-19 generates a substantially higher mortality rate across all or part of the population than would otherwise have been expected, this would support much more radical action to control the pandemic. On the other hand, if COVID-19 has little additional impact on the mortality rate compared to what would otherwise have been expected, then it may be that the negative health impacts of the control measures (for example, due to the impacts of closing down large sectors of the economy^6^) outweigh the positive impact of mitigating the pandemic.

Given the importance of all the above, the aim of this paper is to apply rigorous epidemiological methods using consistent definitions to compare age-sex-standardised mortality and life expectancy contributions of mortality from COVID-19 with socioeconomic inequality and two examples of causes of death which are experienced particularly unequally and which have been the focus of some recent policy attention: suicide and drug-related poisonings.

## Methods

### Estimating COVID-19 mortality

The UK Government has based decisions on pandemic control measures on the modelling of the Imperial College team on the likely scale and timing of the pandemic. The estimate (published on 16^th^ March 2020) of the number of deaths in an unmitigated epidemic in Great Britain (GB) is between 410,000 and 550,000 (with a best estimate of 510,000), and between 5,600 and 48,000 in a fully mitigated epidemic (with a best estimate of 20,000).Estimated deaths by age group are not provided, but ‘infection fatality ratios’ (the percentage of people who are infected who are then expected to die) for 10 year age bands up to age 79 years, with an open upper age band, are provided. Using the 2018 mid-year population data from the Office for National Statistics (ONS) for these age bands for GB and assuming that the proportion of the population in each age group that become infected is uniform, we weighted the deaths in each age group to achieve the total number of deaths estimated by Ferguson et al. We fitted a linear regression model to the logarithm of the infection fatality rates in order to estimate the rate for each 5 year, rather than 10 year, age bands up to age 90 years (Figure S1). These rates were then applied to population data and the total scaled to the estimated deaths in GB under the two scenarios (mitigated and unmitigated).

### Estimating suicide, drug poisoning and inequality-related mortality

We obtained population counts for England, Northern Ireland, Scotland and Wales for each year between 2013 and 2017, by 5 year age band and sex, from the Office for National Statistics (ONS), National Records of Scotland (NRS) and the Northern Ireland Statistics and Research Agency (NISRA). We also obtained the count of all deaths by each nation’s deprivation decile by age and sex, and for two specific causes of death (suicide, including events of undetermined intent; and drug poisoning) by age and sex for each year between 2013 and 2017. The ICD10 codes for suicide were X60-X84, Y10-Y34, Y87.0 and Y87.2; and for drug-related poisonings were F11-F16, F18, F19, X40-X44, X60-X64, X85 and Y10-Y14. The drug poisoning deaths are coded using the broader definition, and the suicide coding uses the older (pre-2011) codes. The definitions of suicide and drug poisoning deaths overlap and cannot therefore be summed.^8^ Following Lewer,^9^ we defined deaths due to inequality as all deaths higher than the rate of mortality in the least deprived tenth of the population in each population. Deprivation was measured using IMD2015 for England, WIMD2014 for Wales, SIMD2016 for Scotland and the 2010 Northern Ireland Multiple Deprivation Measure (NIMDM) for Northern Ireland. We also performed a sensitivity analysis which attributed all deaths above the age-specific rate across every population compared to the least deprived tenth of England (as the nation with the lowest mortality rate in the least deprived tenth).

### Estimating the contribution of different causes to crude and standardised mortality rates, and life expectancy

For crude deaths, we simply summed the reported number of deaths in each age band (taking the 5 year mean between 2013 and 2017 for suicide, drug-and inequality-related deaths). For age-standardised deaths we applied the age-specific deaths to the 2013 European Standard Population to calculate directly standardised estimates. Overall life expectancy was estimated using the method detailed by Auger et al^10^ based on counts of death and populations in 5-year age groups (together with deaths as age 0, 1-4 and aged 90 and over). For COVD-19 and inequality deaths in Northern Ireland we had to estimate the distribution between the 0-1 and 1-4 year age bands as this breakdown was not available. The effect of COVID was estimated by adding the predicted deaths under the two scenarios, re-calculating the life expectancy and taking the difference. The effect of the other causes on life expectancy was estimated by subtracting the deaths attributable to those causes, re-calculating life expectancy and taking the difference.

## Results

Table 1 provides the estimates of the unmitigated and fully mitigated mortality impact of the COVID-19 pandemic by age group for the UK and its nations, based on the Imperial College modelling. It estimates that the majority of deaths will be in the older age groups because the smaller population size is outweighed by the higher mortality rates for these ages. For the UK overall there are estimated to be 195,420 and 7,664 deaths amongst those aged 80+ years in the unmitigated and mitigated scenarios respectively, with 28% of all deaths occurring under the age of 70 years and 38% of all deaths occurring for those aged 80+ years. The proportions are very similar across the nations.

**Table 1.**
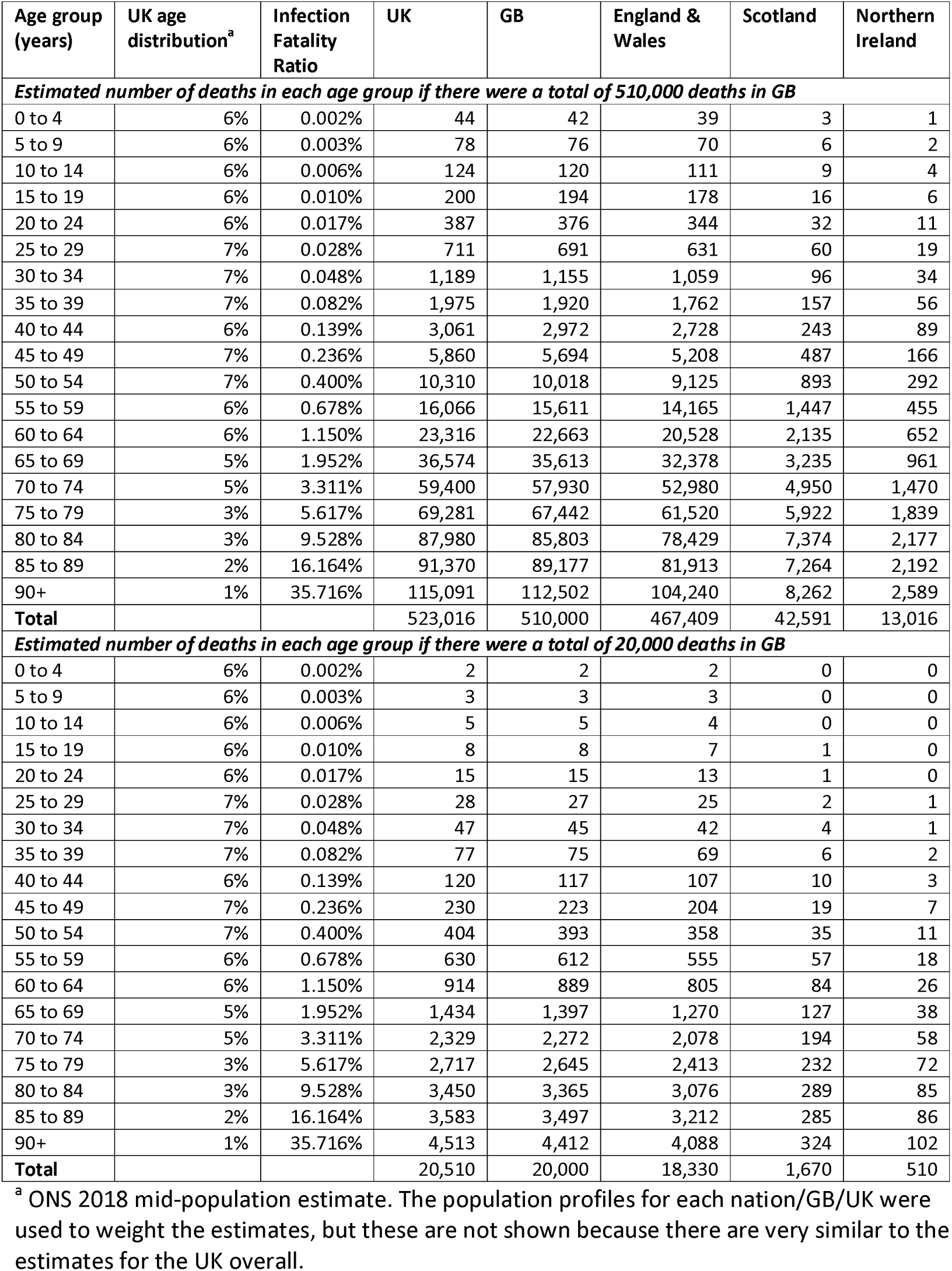
Estimates of the potential mortality impact of COVID-19 on different age groups by applying assumptions to the Imperial model

| Age group (years) | UK age distribution<sup>a</sup> | Infection Fatality Ratio | UK | GB | England & Wales | Scotland | Northern Ireland |
| --- | --- | --- | --- | --- | --- | --- | --- |
| ***Estimated number of deaths in each age group if there were a total of 510,000 deaths in GB*** |  |  |  |  |  |  |  |
| 0 to 4 | 6% | 0.002% | 44 | 42 | 39 | 3 | 1 |
| 5 to 9 | 6% | 0.003% | 78 | 76 | 70 | 6 | 2 |
| 10 to 14 | 6% | 0.006% | 124 | 120 | 111 | 9 | 4 |
| 15 to 19 | 6% | 0.010% | 200 | 194 | 178 | 16 | 6 |
| 20 to 24 | 6% | 0.017% | 387 | 376 | 344 | 32 | 11 |
| 25 to 29 | 7% | 0.028% | 711 | 691 | 631 | 60 | 19 |
| 30 to 34 | 7% | 0.048% | 1,189 | 1,155 | 1,059 | 96 | 34 |
| 35 to 39 | 7% | 0.082% | 1,975 | 1,920 | 1,762 | 157 | 56 |
| 40 to 44 | 6% | 0.139% | 3,061 | 2,972 | 2,728 | 243 | 89 |
| 45 to 49 | 7% | 0.236% | 5,860 | 5,694 | 5,208 | 487 | 166 |
| 50 to 54 | 7% | 0.400% | 10,310 | 10,018 | 9,125 | 893 | 292 |
| 55 to 59 | 6% | 0.678% | 16,066 | 15,611 | 14,165 | 1,447 | 455 |
| 60 to 64 | 6% | 1.150% | 23,316 | 22,663 | 20,528 | 2,135 | 652 |
| 65 to 69 | 5% | 1.952% | 36,574 | 35,613 | 32,378 | 3,235 | 961 |
| 70 to 74 | 5% | 3.311% | 59,400 | 57,930 | 52,980 | 4,950 | 1,470 |
| 75 to 79 | 3% | 5.617% | 69,281 | 67,442 | 61,520 | 5,922 | 1,839 |
| 80 to 84 | 3% | 9.528% | 87,980 | 85,803 | 78,429 | 7,374 | 2,177 |
| 85 to 89 | 2% | 16.164% | 91,370 | 89,177 | 81,913 | 7,264 | 2,192 |
| 90+ | 1% | 35.716% | 115,091 | 112,502 | 104,240 | 8,262 | 2,589 |
| **Total** |  |  | **523,016** | **510,000** | **467,409** | **42,591** | **13,016** |
| ***Estimated number of deaths in each age group if there were a total of 20,000 deaths in GB*** |  |  |  |  |  |  |  |
| 0 to 4 | 6% | 0.002% | 2 | 2 | 2 | 0 | 0 |
| 5 to 9 | 6% | 0.003% | 3 | 3 | 3 | 0 | 0 |
| 10 to 14 | 6% | 0.006% | 5 | 5 | 4 | 0 | 0 |
| 15 to 19 | 6% | 0.010% | 8 | 8 | 7 | 1 | 0 |
| 20 to 24 | 6% | 0.017% | 15 | 15 | 13 | 1 | 0 |
| 25 to 29 | 7% | 0.028% | 28 | 27 | 25 | 2 | 1 |
| 30 to 34 | 7% | 0.048% | 47 | 45 | 42 | 4 | 1 |
| 35 to 39 | 7% | 0.082% | 77 | 75 | 69 | 6 | 2 |
| 40 to 44 | 6% | 0.139% | 120 | 117 | 107 | 10 | 3 |
| 45 to 49 | 7% | 0.236% | 230 | 223 | 204 | 19 | 7 |
| 50 to 54 | 7% | 0.400% | 404 | 393 | 358 | 35 | 11 |
| 55 to 59 | 6% | 0.678% | 630 | 612 | 555 | 57 | 18 |
| 60 to 64 | 6% | 1.150% | 914 | 889 | 805 | 84 | 26 |
| 65 to 69 | 5% | 1.952% | 1,434 | 1,397 | 1,270 | 127 | 38 |
| 70 to 74 | 5% | 3.311% | 2,329 | 2,272 | 2,078 | 194 | 58 |
| 75 to 79 | 3% | 5.617% | 2,717 | 2,645 | 2,413 | 232 | 72 |
| 80 to 84 | 3% | 9.528% | 3,450 | 3,365 | 3,076 | 289 | 85 |
| 85 to 89 | 2% | 16.164% | 3,583 | 3,497 | 3,212 | 285 | 86 |
| 90+ | 1% | 35.716% | 4,513 | 4,412 | 4,088 | 324 | 102 |
| **Total** |  |  | **20,510** | **20,000** | **18,330** | **1,670** | **510** |

<sup>a</sup> ONS 2018 mid-population estimate. The population profiles for each nation/GB/UK were used to weight the estimates, but these are not shown because there are very similar to the estimates for the UK overall.

Table 2 shows the annual (five year mean) age-specific mortality counts for suicide, drug poisoning and inequality-related mortality by nation. These causes have a much younger age distribution of deaths compared to COVID-19, with the highest UK age-specific mortality counts being among 45-49 year olds for suicides and 40-44 year olds for drug poisonings. The peak age for crude mortality due to inequality was 80-84 years but there was a broad age distribution. Suicide and drug poisonings have lower overall counts compared to mitigated COVID-19. However, deaths due to inequality are seven times those of mitigated COVID-19. The crude mortality is substantially higher for men than women for suicide, drug poisonings and inequality (Tables S1 and S2). The modelling of COVID-19 deaths was not available separately by sex. The sensitivity analysis which used the least deprived tenth of the population of England as the comparator increased the number of deaths attributable to inequality slightly as the least deprived tenth of England has lower mortality than the equivalents across the other nations. However, the impact on the overall results is small (Table S3).

**Table 2.** Estimated number of deaths due to suicide, drug poisonings and inequality-related deaths by age (2013-2017 annualised mean, total population)

| Age group (years) | Drug poisonings |  |  |  |  | Suicide |  |  |  |  | Inequality |  |  |  |  |
| --- | --- | --- | --- | --- | --- | --- | --- | --- | --- | --- | --- | --- | --- | --- | --- |
|  | UK | GB | E&W | Sco | NI | UK | GB | E&W | Sco | NI | UK | GB | E&W | Sco | NI |
| 0-4 | 1 | 1 | 1 | 0 | 0 | 4 | 4 | 3 | 1 | 0 | 1,040 | 1,043 | 983 | 60 | -3 |
| 5-9 | 1 | 1 | 1 | 0 | 0 | 1 | 1 | 1 | 0 | 0 | 97 | 93 | 92 | 1 | 4 |
| 10-14 | 3 | 3 | 2 | 1 | 0 | 20 | 18 | 15 | 3 | 2 | 58 | 54 | 45 | 9 | 4 |
| 15-19 | 63 | 59 | 46 | 13 | 4 | 195 | 182 | 161 | 20 | 14 | 280 | 255 | 222 | 33 | 25 |
| 20- 24 | 194 | 181 | 150 | 31 | 13 | 395 | 364 | 321 | 43 | 31 | 266 | 221 | 152 | 69 | 46 |
| 25-29 | 356 | 339 | 271 | 67 | 17 | 461 | 428 | 370 | 58 | 33 | 583 | 562 | 440 | 122 | 21 |
| 30-34 | 528 | 508 | 395 | 114 | 19 | 519 | 483 | 422 | 61 | 36 | 1,160 | 1,122 | 890 | 232 | 38 |
| 35-39 | 672 | 655 | 516 | 139 | 17 | 531 | 500 | 432 | 68 | 31 | 1,958 | 1,880 | 1,557 | 324 | 78 |
| 40-44 | 734 | 719 | 558 | 161 | 15 | 663 | 633 | 547 | 86 | 31 | 3,189 | 3,119 | 2,651 | 467 | 70 |
| 45-49 | 632 | 618 | 490 | 127 | 14 | 737 | 703 | 611 | 92 | 34 | 5,032 | 4,853 | 4,226 | 627 | 179 |
| 50-54 | 467 | 456 | 365 | 91 | 11 | 684 | 653 | 574 | 80 | 31 | 6,470 | 6,287 | 5,412 | 876 | 183 |
| 55-59 | 284 | 277 | 232 | 46 | 6 | 523 | 504 | 442 | 62 | 19 | 8,377 | 8,149 | 6,993 | 1,156 | 228 |
| 60-64 | 189 | 184 | 158 | 26 | 5 | 365 | 350 | 309 | 41 | 15 | 11,473 | 11,197 | 9,707 | 1,490 | 276 |
| 65-69 | 116 | 115 | 100 | 15 | 2 | 287 | 278 | 249 | 29 | 9 | 16,053 | 15,629 | 13,609 | 2,019 | 424 |
| 70-74 | 74 | 72 | 63 | 9 | 2 | 209 | 204 | 180 | 23 | 6 | 18,497 | 18,070 | 15,649 | 2,421 | 426 |
| 75-79 | 56 | 55 | 49 | 6 | 1 | 162 | 158 | 142 | 16 | 4 | 21,398 | 20,961 | 18,371 | 2,591 | 436 |
| 80-84 | 41 | 40 | 39 | 2 | 1 | 128 | 127 | 118 | 9 | 1 | 22,305 | 21,982 | 19,501 | 2,481 | 322 |
| 85- 89 | 29 | 29 | 27 | 2 | 0 | 95 | 94 | 88 | 5 | 1 | 17,154 | 16,886 | 15,194 | 1,692 | 268 |
| 90+ | 22 | 22 | 21 | 1 | 0 | 56 | 56 | 52 | 4 | 0 | 11,957 | 11,799 | 11,319 | 481 | 158 |
| **Total** | 4,460 | 4,334 | 3,483 | 850 | 126 | 6,038 | 5,739 | 5,039 | 701 | 298 | 147,346 | 144,164 | 127,013 | 17,150 | 3,182 |

Notes: UK = United Kingdom; GB = Great Britain; E&W = England & Wales; Sco = Scotland; NI = Northern Ireland

Suicide, drug poisonings and inequalities all generate mortality every year, and cumulatively over a few years amount to greater total crude deaths than COVID-19. For example, in 3.5 years the total death count for inequality-related deaths overtakes the number of COVID-19 deaths in the UK in the completely unmitigated scenario.

Table 3 compares the crude mortality, age-standardised mortality and life expectancy contributions of the different causes. The negative contribution to life expectancy for drug, suicide and inequality related deaths is much greater than the contribution from the fully-mitigated COVID-19 mortality. The contribution to life expectancy of inequality-related deaths is higher, and the contribution of COVID-19 deaths lower, in Scotland and Northern Ireland compared to the UK overall.

**Table 3.** Comparison of the impacts of COVID and other causes on mortality indices

| **Age group (years)** | **UK** | **GB** | **England & Wales** | **Scotland** | **Northern Ireland** |
| --- | --- | --- | --- | --- | --- |
| ***Crude deaths*** |  |  |  |  |  |
| COVID-19 unmitigated | 523,016 | 510,000 | 467,409 | 42,591 | 13,016 |
| COVID-19 fully mitigated | 20,510 | 20,000 | 18,330 | 1,670 | 510 |
| Drug poisonings | 4,460 | 4,334 | 3,483 | 850 | 126 |
| Suicide | 6,038 | 5,739 | 5,039 | 701 | 298 |
| Inequality-related | 147,346 | 144,164 | 127,013 | 17,150 | 3,182 |
| ***Age-standardised mortality per 100,000*** |  |  |  |  |  |
| COVID-19 unmitigated | 903 | 903 | 903 | 899 | 910 |
| COVID-19 fully mitigated | 35 | 35 | 35 | 35 | 36 |
| Drug poisonings | 7 | 7 | 6 | 16 | 7 |
| Suicide | 9 | 9 | 9 | 13 | 16 |
| Inequality-related | 253 | 254 | 245 | 346 | 210 |
| ***Impact on life expectancy (years)*** |  |  |  |  |  |
| COVID-19 unmitigated | -5.96 | -5.96 | -6.03 | -5.26 | -5.76 |
| COVID-19 fully mitigated | -0.33 | -0.33 | -0.34 | -0.29 | -0.32 |
| Drug poisonings | -0.20 | -0.20 | -0.17 | -0.45 | -0.22 |
| Suicide | -0.25 | -0.25 | -0.24 | -0.34 | -0.49 |
| Inequality-related | -3.51 | -3.53 | -3.40 | -4.73 | -3.02 |

The lower age distribution of suicide, drug poisoning and inequality-related deaths reduces further the difference between them and unmitigated COVID-19 when using the age-standardised and life expectancy contribution measures, such that the negative contribution to life expectancy of unmitigated COVID-19 represents the equivalent of the inequality-related deaths occurring over only 1.7 years for the UK overall. This means that over 10 years there is a greater negative life expectancy contribution from inequality than about six unmitigated COVID-19 pandemics for the UK overall.

## Discussion

### Main results

COVID-19 represents a large and urgent mortality risk for populations across the world. However, counting cumulative crude deaths is an unhelpful means of ascertaining the scale of this risk. Comparing standardised mortality rates and life expectancy contributions of COVID-19 and three causes of death that are strongly socially determined - suicide, drug poisoning and inequality-related deaths – reveals that the mortality from a fully unmitigated COVID-19 pandemic is modelled to be responsible for the negative life expectancy contribution that occurs due to the cumulative inequality-related deaths over the course of around 1.7 years. Putting this another way, over a ten year period if there were around six unmitigated pandemics on the scale of the current COVID-19 pandemic, the impact on life expectancy would be less than that of inequalities.

### Strengths and limitations

There is an urgency in being able to calibrate the mortality risk due to COVID-19 in order to be able to ascertain the appropriate level of control measures required. This study therefore uses the available modelled data that have been used to justify the control measures in place at the time of writing alongside routinely available and published data on other mortality risks that have been present in the UK and around the world for some time.^11^ There are, however, a number of limitations. We have had to model the age-specific mortality of COVID-19 by 5 year age band as this was not available in the Imperial College data. The exponential relationship between age and COVID-19 mortality alongside the open upper age bound in our data make the estimates at this upper age more uncertain. Similarly, the Imperial College modelling does not provide estimates by sex, although the emerging evidence suggests that the mortality rates are higher amongst men. This means that the life expectancy impacts of COVID-19 provided in this paper are likely to be overestimated because men have a systematically lower life expectancy than women and so the loss of lifespan for men will be less. It is also recognised that COVID-19 mortality rates are higher amongst those with co-morbidities for any given age-sex group.^12^ Our modelling does not differentiate between groups on this basis and is therefore likely to be a further source of systematic bias which overestimates the mortality impact of COVID-19. For these reasons, the impacts on mortality of COVID-19 estimated here are likely to be substantially higher than reality.

### How this fits with the existing literature

The COVID-19 pandemic and health inequality both clearly require a radical policy response to reduce the associated mortality. Health inequalities have long been recognised as an important policy issue^11,13^ but there has been little or no progress at reducing them over at least 40 years.^14^ For COVID-19, it is also important to ensure that the social distancing measures (the substantive difference between an unmitigated and fully mitigated COVID-19 pandemic) do not cause more population health damage than the gain achieved through mitigation.^15^ Social distancing is likely to have marked impacts on the economy, incomes, employment, social isolation, physical activity and education. A rapid Health Impact Assessment has identified that the potential for unintended negative health consequences is very large.^16^ Although many of the relevant mechanisms and negative impacts have been reduced to some extent in the UK and elsewhere (e.g. through the policy of funding part of the wages of staff who have been furloughed), it is highly unlikely that these can be reduced to zero, particularly given the scale of the economic shock.^17^

### Implications

The policy response to public health challenges should match the mortality risk. The analysis in this paper indicates that the long-term life expectancy impact of inequalities is substantially greater than even an unmitigated COVID-19 pandemic because the problem of inequalities is ongoing. The rapid policy response to COVID-19 demonstrates what governments can and should do in the face of a massive population health challenge. Yet the mortality risk from the socially-generated causes compared here, as well as many others, over only a few short years contributes many more deaths on all metrics than COVID-19. It is interesting to compare the radical government action in the face of the COVID-19 threat but much less drastic policy interventions to reduce income, wealth and power inequalities (e.g. through social security benefit values, progressive taxes, ownership of capital, etc.^13,18,19^) to reduce inequality-related mortality. The post-COVID-19 pandemic period should be used to ‘build back better’ and ensure that society and the economy in the future provides the basis to reduce social inequalities in health and all avoidable causes of death.^6^ Future monitoring and reporting of COVID-19 mortality should include age-standardised mortality rates and life expectancy contributions for set time periods rather than simply reporting cumulative crude deaths. The estimation of the life expectancy contribution of COVID-19 should also be repeated later in the pandemic when actual (rather than modelled) mortality data are available for the population overall and stratified by sex.

## Conclusion

The mortality risk from COVID-19 is substantial and if unmitigated could lead to a decline of 5.96 years of life expectancy. The risks from recurrent deaths, such as those due to inequality, will quickly surpass those due to COVID-19. Building the economy back better such that inequality-related deaths are reduced in the future is as important as social distancing is now for future population health.

## Data Availability

We do not own the data used to undertake the analysis in this paper but they are available on request from ONS, NRS and NISRA.

## Competing interests

The authors declare that they have no competing interests

## Notes

### Competing Interest Statement

The authors have declared no competing interest.

### Funding Statement

RD and AHL are funded by the Medical Research Council (MC_UU_12017/13) and the Chief Scientist Office (SPHSU13).

